# Forecasting the Epidemiological Impact of Coronavirus Disease (COVID-19): Pre-vaccination Era

**DOI:** 10.1101/2021.03.17.21253791

**Authors:** Saheed Oladele Amusat

**Affiliations:** Federal Medical Centre Birnin Kudu, Jigawa State, Nigeria; College of Health Sciences, Ladoke Akintola University of Technology Osogbo, Nigeria

**Keywords:** Pandemic, Epidemiology, COVID-19, Coronavirus, nCOV

## Abstract

**Background:** During this pandemic, many studies have been published on the virology, diagnosis, prevention, and control of the novel coronavirus. However, fewer studies are currently available on the quantitative future epidemiological impacts. Therefore, the purpose of this study is to forecast the COVID-19 morbidities and associated-mortalities among the top 20 countries with the highest number of confirmed COVID-19 cases globally prior to vaccination intervention.

**Method:** We conducted a secondary data analysis of the prospective geographic distribution of COVID-19 cases data worldwide as of 10 April 2020. The historical data was forecasted using Exponential-Smoothing to detect seasonality patterns and confidence intervals surrounding each predicted value in which 95 percent of the future points are expected to fall based on the forecast.

**Results:** The total mean forecasted cases and deaths were 99,823 and 8,801. Interestingly, the US has the highest forecasted cases, deaths, and percentage cases-deaths ratio of 45,338, 2 358, and 5.20% respectively. China has the lowest cases, deaths, and percentage cases-deaths ratio −267, −2, and 0.75% respectively. In addition, France has the highest forecasted percentage cases-deaths ratio of 26.40% with forecasted cases, and deaths of 6,246, and 1,649 respectively.

**Conclusion:** Our study revealed the possibility of higher COVID-19 morbidities and associated-mortalities worldwide.

## BACKGROUND

The COVID-19 pandemic is the most impromptu global health emergency of the 21st century. Coronavirus disease (COVID-19) represents a potentially fatal disease that is of great global public health concern (1). Coronaviruses may cause various symptoms ranges from acute pneumonia, fever, difficulty breathing, and acute pulmonary infection (2). These viruses are common in animals worldwide, but very few cases have been known to affect humans (3). The World Health Organization (WHO) adopted the term 2019-novel coronavirus (2019-nCOV) to refer to a specie of coronaviruses that affected the lower respiratory tract of pneumonia patients in Wuhan, Hubei province, China on 29 December 2019 (4,5,6). The WHO officially renamed the 2019 novel coronavirus has coronavirus disease (COVID-19) which is now universally being used (WHO, 2020). In the same vein, the current reference name for the virus is Severe Acute Respiratory Syndrome coronavirus 2 (SARS-CoV-2). It was reported that a cluster of patients with pneumonia of unknown etiology was linked to a local Huanan South China wet-Market in Wuhan, in December 2019 (7).

According to ECDC (2020), over 1 316 988 cases of COVID-19 have been reported worldwide, with more than 74,066 deaths since 31 December 2019, and as of 7 April 2020, approximately 0.5% of these cases (608 500) have been reported from the European Union, European Economic Area (EEA) countries and the UK and approximately 0.1% (51 059) of them have died (8). Also, and recently, the European all-cause mortality monitoring system showed all-cause excess mortality above the expected rate in Belgium, France, Italy, Malta, Spain, Switzerland, and the United Kingdom, mainly in the age group of 65 years and above (9).

Based on the Center for Disease Control (2020), community transmission of COVID-19 was first reported in the United States in February 2020. By the middle of March, all the 52 states and four U.S. territories had reported cases of COVID-19. As of April 7, 2020, a total of 395,926 COVID-19 cases, including 12,757 deaths, were reported in the United States. In place of these developments, advanced countries are experiencing a dramatic impact on the epidemic. Thus, this study aims to forecast the future impact of COVID-19 on the 20 countries with the highest number of confirmed cases in the ongoing pandemic.

## METHODOLOGY

We conducted a secondary data analysis of the prospective geographic distribution of COVID-19 cases data worldwide as of 10 April 2020. The data was sourced from the European Centre for Disease Prevention and Control, which was collected by the ECDC’s Intelligence team based on reports from global healthcare authority. According to ECDC, there are regular updates from European Union and EEA countries through the Early Warning and Response System (EWRS), The European Surveillance System (TESSy), the World Health Organization (WHO), and email exchanges with other international stakeholders.

### Data analysis

The historical data was forecasted using Exponential Smoothing to detect seasonality patterns and confidence intervals surrounding each predicted value in which 95 percent of the future points are expected to fall based on the forecast. For convenient interpretation, the mean forecast, the confidence intervals of cases, and deaths for each country were recorded (Table 1). Furthermore, the percentage death-case ratio was reported to determine the degree of mortality in each country (Figure 3) by determining the forecasted mean to deaths multiplied by 100.

**Table 1:**
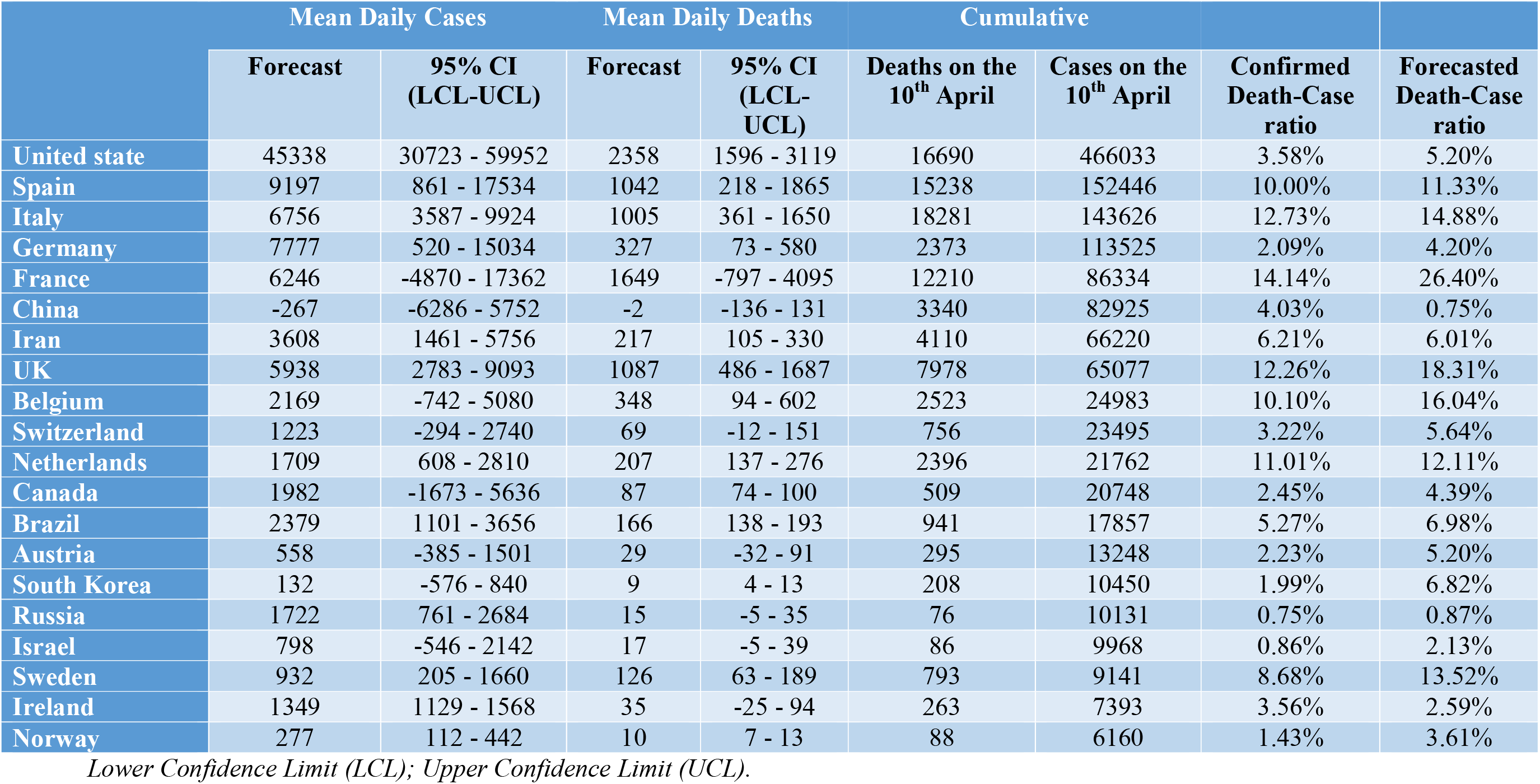
The mean forecast statistics and death-case ratio among the countries.

## RESULTS AND DISCUSSION OF FINDINGS

The historical COVID-19 data of the top 20 countries with the highest number of confirmed COVID-19 cases from 31 December 2019 to 10 April 2020 was forecasted for 103 days starting from 10 April to 21 July 2020. As can be seen in table 1, the US has the highest number of confirmed cases 466,033 while Norway has the lowest number of confirmed cases 6,160 among the countries studied as of 10 April 2020. The US has the highest forecasted mean daily confirmed cases, and deaths of 45,338; CI (30,723-59,952) and 2,538; CI (1,596-3,119) respectively (Table 1 and Figure 1). The mean deaths-cases ratio was 3.58% as of 10 April 2020 while the forecasted mean deaths-cases ratio was 5.20%. This implies that the US could experience more morbidities and associated-mortalities taking into account the onset of community transmission, and time of the study. Interestingly, China has the lowest forecasted mean daily cases and deaths of −267; CI (−6286-5762), and −2; CI (−136-131) respectively (Table 1). Although, the deaths-cases ratio was 4.03% as of 10 April 2020, and the forecasted deaths-cases ratio was 0.75%. Similarly, the mean deaths-cases ratio in, Ireland, and Iran were predicted to be 2.59%, and 6.01% compared to their actual deaths-cases as of 10 April 2020 (Figure 2 and Table 1). This implies that there is a high tendency that China, Ireland, and Iran could record lower COVID-19 associated death(s) during the forecasted term. Although this depends on how compliant people are practicing protective measures and how effective the implemented protective policies. After the US, Spain, Germany, Italy, France and the UK were the top 5 countries with the highest forecasted mean daily cases of 9,197; CI (861-17534), 7,777; CI (520-15034), 6,756; CI (3587-9924), 6,246; CI (−4870-17362), and 5,938; CI (2783-9093) respectively. However, France, UK, Spain, Italy, and Belgium have the highest forecasted mean daily deaths of 1,649; CI (−797-4095), 1,087; CI (486-1687), 1,042; CI (218-1865), 1,005; CI (361-1650), and 348; CI (94-602) respectively after the USA (Table 1). The mean deaths-cases ratio showed that France had the highest of 14.14% while Russia had the lowest mean death-case ratio of 0.75% as of 10 April 2020. Followed after France is Italy, UK, Netherlands, Belgium, Spain, and Sweden have the highest incidences with mean death-case ratio of 12.73%, 12.26%, 11.01%, 10.10%, 10.0%, and 8.68% respectively (Figure 3) as of 10 April 2020. Also, the forecasted death-case ratio showed that France, the UK, Belgium, Italy, Spain, Netherlands and Sweden could be more vulnerable to COVID-19 associated-mortality during the forecasted period with the mean forecasted deaths-cases ratio of 26.40%, 18.31%, 16.04%, 14.88%, 11.33%, 12.11% and 13.52% respectively. Lastly, the overall results showed that increase COVID-19 cases and associated-death were very much likely to occur in the countries.

**Figure 1:**
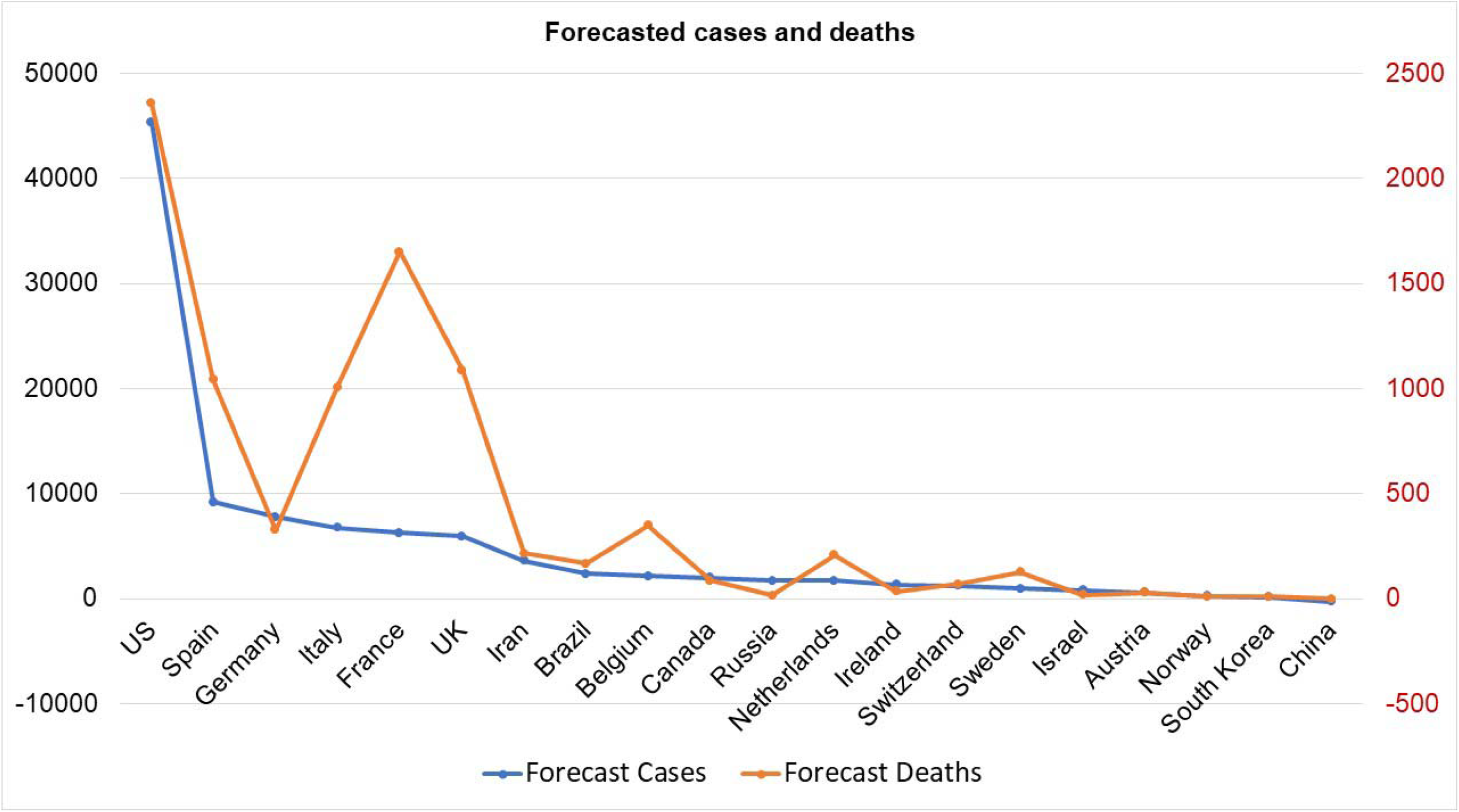
The chart showing the mean forecasted cases and deaths.

**Figure 2:**
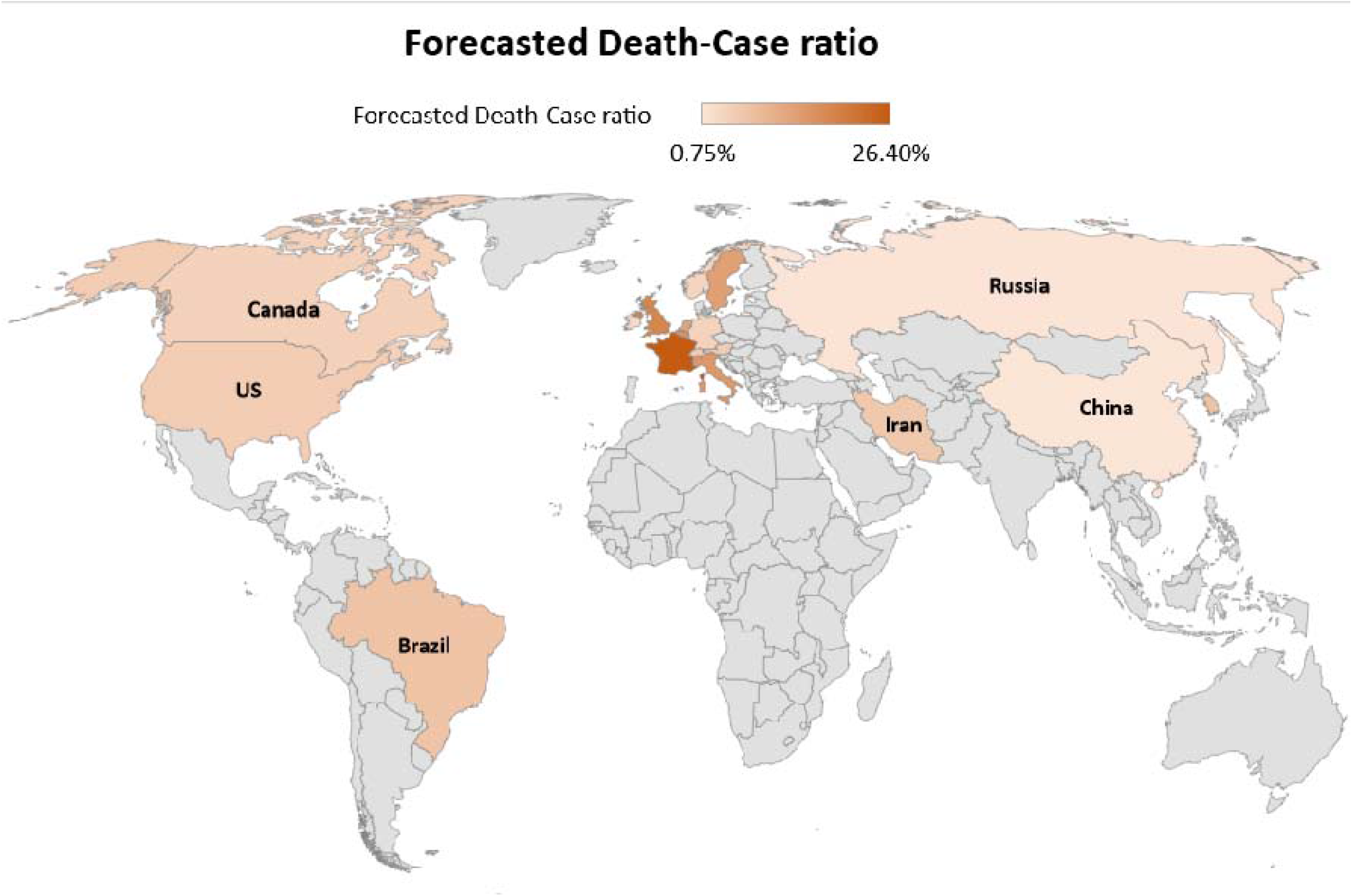
The world map showing the forecasted death–case ratio in countries studied.

**Figure 3:**
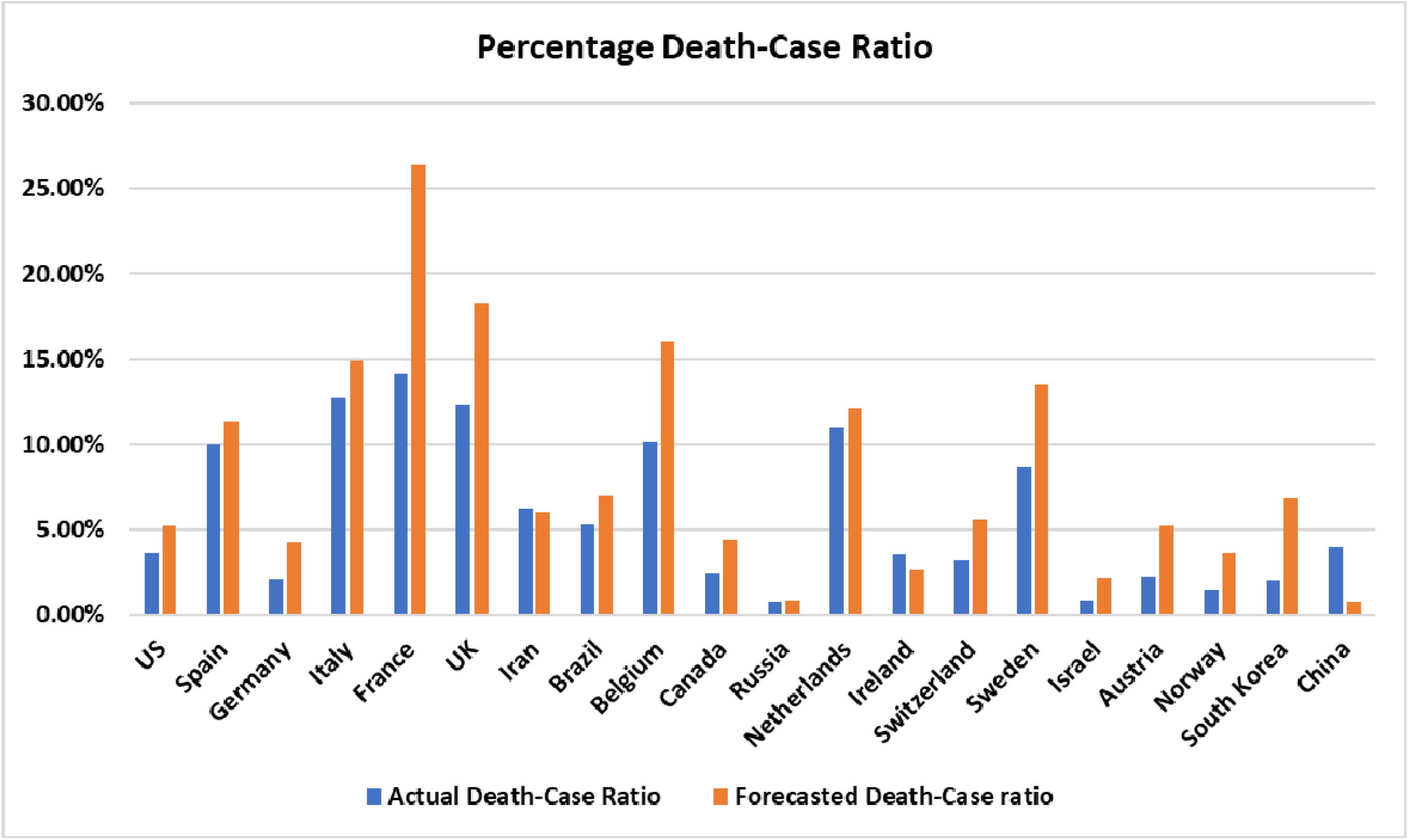
The comparative chart showing the actual, and forecasted death-case ratio.

### Strengths and limitations of the study

The study employed a systematic and rigorous approach to source for the information relevant to these research objectives. This research summarizes global distributions and quantitative impacts of COVID-19 on global health. This will stimulate the research community, stakeholders in the government and non-governmental organizations, and healthcare professionals to optimize strategies to mitigate the impact of the ongoing pandemic and strengthening preparedness for any future outbreaks. Also, it will inform the entire public to emulate compliance towards personal protective measures against the future outbreak. Our study only focuses on the top 20 countries with the highest confirmed cases in the COVID-19 global timeline. Although, it failed to reflect the entire COVID-19 cases worldwide due to redundancy in some countries’ figures.

## CONCLUSION

Our study showed a holistic view of the current research in response to the outbreak of COVID-19. During this pandemic, many studies have been published on the virology, causes, clinical manifestation and diagnosis, and prevention and control of the novel coronavirus. However, fewer studies are currently available on the quantitative impact of COVID-19 both in the present-term and long term. Research in this aspect is urgently needed to inform stakeholders at local, regional, national, and global levels to devise strategies towards containment of SARS-COV-2 and mitigation of the impact of the outbreak on global health and economy. We suggest further research to be conducted at local, regional, and global levels to provide valid and reliable ways to manage this kind of public health emergency in both the short-term and long-term. Also, accurate and timely infectious disease forecasts could inform stakeholders and authorities’ responses to both future epidemics and pandemics by providing strategic policies for preparedness towards prevention and mitigation. Despite current limitations, forecasting remained a powerful tool to aid public health decision-making.

## Data Availability

The data is openly accessible via European Centre for Disease Prevention and Control

## ACKNOWLEDGMENT

We thank the European Centre for Disease Prevention and Control for making the geographic distribution of COVID-19 cases data available and accessible through https://www.ecdc.europa.eu/en/novel-coronavirus-china.

